# SARS-CoV-2 testing in the Slovak Republic from March 2020 to September 2022 – summary of the pandemic trends

**DOI:** 10.1101/2023.06.26.23291891

**Authors:** Nikola Janostiakova, Andrej Gnip, Dominik Kodada, Rami Saade, Gabriela Blandova, Emilia Mikova, Elena Tibenska, Vanda Repiska, Gabriel Minarik

**Affiliations:** Comenius University in Bratislava Faculty of Medicine, Department of Clinical Biology, Genetics and Clinical Genetics, Bratislava, Slovakia; Medirex, a.s., Pezinok, Slovakia; Medirex Group Academy n.p.o., Nitra, Slovakia

**Keywords:** COVID-19, pandemic, Slovakia, statistics, RT-qPCR, Ct value, age groups

## Abstract

The COVID-19 pandemic has been part of Slovakia since March 2020. Intensive laboratory testing ended in October 2022, when the number of tests dropped significantly, but the state of the pandemic continues to this day. For the management of COVID-19, it is important to find an indicator that can predict pandemic changes in the community. The average daily/weekly Ct value with a certain time delay can predict changes in the number of cases of SARS-CoV-2 infection, which can be a useful indicator for the healthcare system. The study analyzed the results of 1,420,572 RT-qPCR tests provided by one accredited laboratory during the ongoing pandemic in Slovakia from March 2020 to September 2022. The total positivity of the analyzed tests was 24.64%. The average Ct values found were the highest in the age group of 3-5 years, equal to the number 30.75; the lowest were in the age group > 65 years, equal to the number 27. The average weekly Ct values ranged from 22.33 (pandemic wave week) to 30.12 (summer week). We have summarized the results of SARS-CoV-2 diagnostic testing in Slovakia with the scope defined by the rate and positivity of tests carried out at Medirex a.s. laboratories.

## 1 Introduction

The COVID-19 pandemic in Slovakia is part of the global pandemic of the disease COVID-19 caused by the SARS-CoV-2 virus. The first case was confirmed in Slovakia on March 6, 2020 [1]. The first measures against the pandemic were also adopted on March 6, 2020 [2]. A state of emergency was first declared on March 12, 2020 [3]. The state of the pandemic continues worldwide to this day [4].

Strict rules and measures adopted in Slovakia at the beginning of the pandemic kept the number of infected people relatively under control. The next wave of the pandemic during the winter of 2020/2021 brought a significant worsening of the situation [5]. Due to the appearance of a new alpha mutation (B.1.1.7), this deterioration was even beyond the expectations of experts [6]. In February 2021, Slovakia became the worst in the world in the number of deaths and hospitalized cases per capita [7]. The inability to predict the rate of change in the SARS-CoV-2 infection hindered the ability to effectively respond to the crisis not only in Slovakia, but also in the world. The possible way how to monitor disease spread and estimate the evolution of pandemic waves is to perform large scale laboratory as well as home-based testing for the SARS-CoV-2 infection in the population and this was adopted in our region. The most widely used laboratory test method for detecting the presence of the SARS-CoV-2 virus in a sample is RT-qPCR (Quantitative Real-Time Reverse Transcription Polymerase Chain Reaction). At November 22, 2022, Slovakia recorded a total of 1,855,129 positive cases of coronavirus from 7,348,616 RT-qPCR tests performed (1,345,853 tests per 1 million inhabitants) [8].

RT-qPCR detects and quantifies the monitored section of DNA in real time reaction. Amplification monitoring is based on the principle of fluorescence using probes (fluorescent substances) that bind specifically or non-specifically to the amplified DNA. An amplification curve is created for each examined sample based on the measured fluorescence signals [9]. For each amplification curve is then determined a threshold value, referred as the Ct (threshold cycle), that is the intersection between an amplification curve and a threshold line and determines the positivity of the test. The Ct value represents the number of PCR cycles required for the fluorescence signal generated by the incorporation of fluorescent dyes into the PCR product to exceed the background fluorescence level [10]. It is a relative measure of the concentration of target in the PCR reaction. Low Ct values indicate a high viral load and a potentially high level of infectivity. High Ct values indicate a very early or late stage of infection, and therefore a potentially low level of infectivity [11].

For the surveillance of COVID-19, it is important to find a good indicator that can predict the trends of the pandemic in the community. The results of several studies indicate that the average daily/weekly Ct value with a certain time delay can predict the increase in the number of positive confirmed cases of COVID-19, which can be a useful indicator for the healthcare system [12, 13]. It is extremely important in diseases, which have a high potential to significantly increase the demand for hospitalization and cause an overcrowding of hospital capacities, threatening a standardly functioning healthcare system For a more effective and sustainable management of the pandemic, it is necessary to predict the improvement/deterioration of the epidemiological situation and prevent its deterioration by following a set of epidemiological criteria at the regional, national, and international level, so that measures are tightened and loosened based on objective criteria [14]. The aim of the work was to summarize the results of SARS-CoV-2 diagnostic testing in Slovakia at the regional level with the scope defined by the rate of tests carried out in Medirex a.s. laboratories and with reference to the course of the pandemic evaluated at the national level. The number of this study data is robust, representing more than 1.4 million evaluated RT-qPCR tests. We made sure that the selection of the date period included the entire timeline of the massive national testing of positivity/negativity of Covid-19 within the Slovak Republic. We were particularly interested in individual parameters in terms of differences in viral load over time and between characterized age subgroups (estimated using Ct values). Our findings regarding the trends of the pandemic in the Slovak population can be used for a more effective setting of either diagnostics or the logistics of measures in possible future epidemics/pandemics.

## 2 Article type

Original Research

## 3 Manuscript Formatting

### 3.1 Materials and methods

The study analyzed data provided by the accredited laboratory Medirex. The data represent the results of RT-qPCR tests of patients and general population. Samples were evaluated from March 2020 to September 2022 and tests were performed in the Central Laboratories in Bratislava, Kosice and Nitra. During this period, the laboratories analyzed 1,420,572 tests. The samples were collected from 74 (93.7%) districts of Slovakia. The representation of the districts in the study is connected with the location of the sample collection points created during the SARS-CoV-2 pandemic testing and the location of the diagnostic laboratories Medirex, a.s. In the case that the sample collection points of certain districts of Slovakia were contracted another diagnostic laboratory, we did not have samples from this district.

Laboratory Medirex, a.s. carried out tests for the general population and for patients based on a request from a doctor as one of the state-contracted diagnostic laboratories in Slovakia. On the website www.korona.gov.sk, citizens had the opportunity to request a SARS-CoV-2 detection test. The Ministry of Health of the Slovak Republic recommended to use the possibility of testing with PCR tests in case of suspicion of the disease COVID-19. If symptoms of infection were entered (e. g. temperature above 38 degrees, dry cough, joint and muscle pain, shivering, coughing or loss of smell or taste) or contact with a positively tested person, this testing was fully covered by public health insurance. Based on the assessment of clinical symptoms, citizens were automatically assigned the nearest possible testing date in the closest possible vicinity of the address they entered in the application. A positive antigen test of citizen was another possibility to be tested similarly with the PCR test - citizens could also verify positivity of antigen test by subsequently performing a PCR test, which in this case was also covered by public health insurance. In addition to free testing in case of symptoms of the SARS-CoV-2 infection, citizens had the opportunity to be tested free of charge with PCR tests during pre-operative examinations or before planned hospitalization [15]. Finally, individuals could undergo the PCR examination also as self-payers, in this case no selection of individuals (assessment of the presence of symptoms, etc.) was necessary to perform the test. The presence of SARS-CoV-2 was determined from nasopharyngeal swabs and saliva samples, since the collection sites in Slovakia provided these two options for collecting genetic material. Automated extraction of nucleic acid (DNA/RNA) on magnetic particles was performed using the Sera-Xtracta Virus/Pathogen Kit (Cytiva) and the Zybio Nucleic Acid Extraction Kit (Zybio) using the KingFisher™ Flex Purification System (Thermo Scientific) and Zybio EXM 3000 Nucleic Acid Isolation System (Zybio) / Zybio EXM 6000 Nucleic Acid Isolation System (Zybio), respectively. Testing was performed by RT-qPCR using the COVID-19 Real Time Multiplex RT-PCR Kit (Lab- systems Diagnostics), SARS-CoV-2 Nucleic Acid Detection Kit (Zybio) and Novel Coronavirus (2019-nCoV) Real Time Multiplex RT-PCR Kit (Liferiver) on qPCR platforms ABI 7500 (Fast) Re- al-Time PCR System, QuantStudio 5 and QuantStudio 6 Real-Time PCR System (Applied Biosystems). Data obtained for each test included daily sample number, collection date, patient ID, age, sex, location of sample collection, test result and, in the case of a positive result, the Ct value of the viral gene E – diagnostic value up to 40 (for the SARS-CoV-2 Nucleic kit Acid Detection Kit (Zybio)) or 41 (for COVID-19 Real Time Multiplex RT-PCR Kit (Labsystems Diagnostics) and Novel Coronavirus (2019-nCoV) Real Time Multiplex RT-PCR Kit (Liferiver)) – that mean 39 or 40 of amplification cycles is needed to reach the threshold at which a molecular diagnostic test can be detected with positive signal. Ct values have been reported and archived for a positive result since 2020-12-03 - therefore, results for Ct values include the date range December 2020 - September 2022.

Data were categorized into nine groups by age/level of school attended: newborns and toddlers (0-2 years); children of preschool age (3-5 years); children from primary school I (6-8 years old); children of primary school II (9-13 years); children of primary school III (14-15 years); youth I/secondary schools (16-19 years); youth II/university (20-26 years); adults (27-65 years) and sen-iors (66 years and older). For the purposes of demographic analysis, the data were also categorized into individual districts/regions of Slovakia. Population data were obtained from census done by Statistical Office of the Slovak Republic in 2021 [16].

Various statistical tests were used to compare categories of data, check for correlations and data characteristics. Fisher’s exact test was used to compare values of positivity and Welch’s t-test was used to compare Ct values between different groups. Hartigans’ dip test was used to check for unimodality of data and Spearman’s rank correlation coefficient was used to address possible correlations. Together 133 statistical tests were performed and Bonferroni correction was used to adjust the threshold of significance for p-value. Thus 3.76E-4 was used as the threshold instead of 0.05.

### 3.2 Results

From March 2020 to September 2022, the laboratory performed a total of 1,420,572 tests for diagnostic purposes, which for the given period represents more than 19% of the total number of RT-qPCR tests in Slovakia. To estimate how the representation of RT-qPCR tests in our study changed over time, we determined the percentage representation of Medirex tests in relation to the total number of RT-qPCR tests in Slovakia on the last day of each month - the representation of tests evaluated in Medirex, a.s. ranged from 8.72% (2022-08-31) to 57.34% (2021-01-31). The population source of the studied population divided into the districts of Slovakia is shown in Figure 1. The color scale shows the number of tests performed and positivity in individual districts. The total positivity of the analyzed tests was 24.64% (350,067 tests), while the highest rate of positivity (over 30%) was achieved by the age categories: 6-8 years, 9-13 years, 14-15 years, 16-19 years. The age distribution of the tested patients of the study, together with the frequency of positive/negative PCR tests and the corresponding rate of positivity for individual age categories, is shown in Table 1.

**Figure 1:**
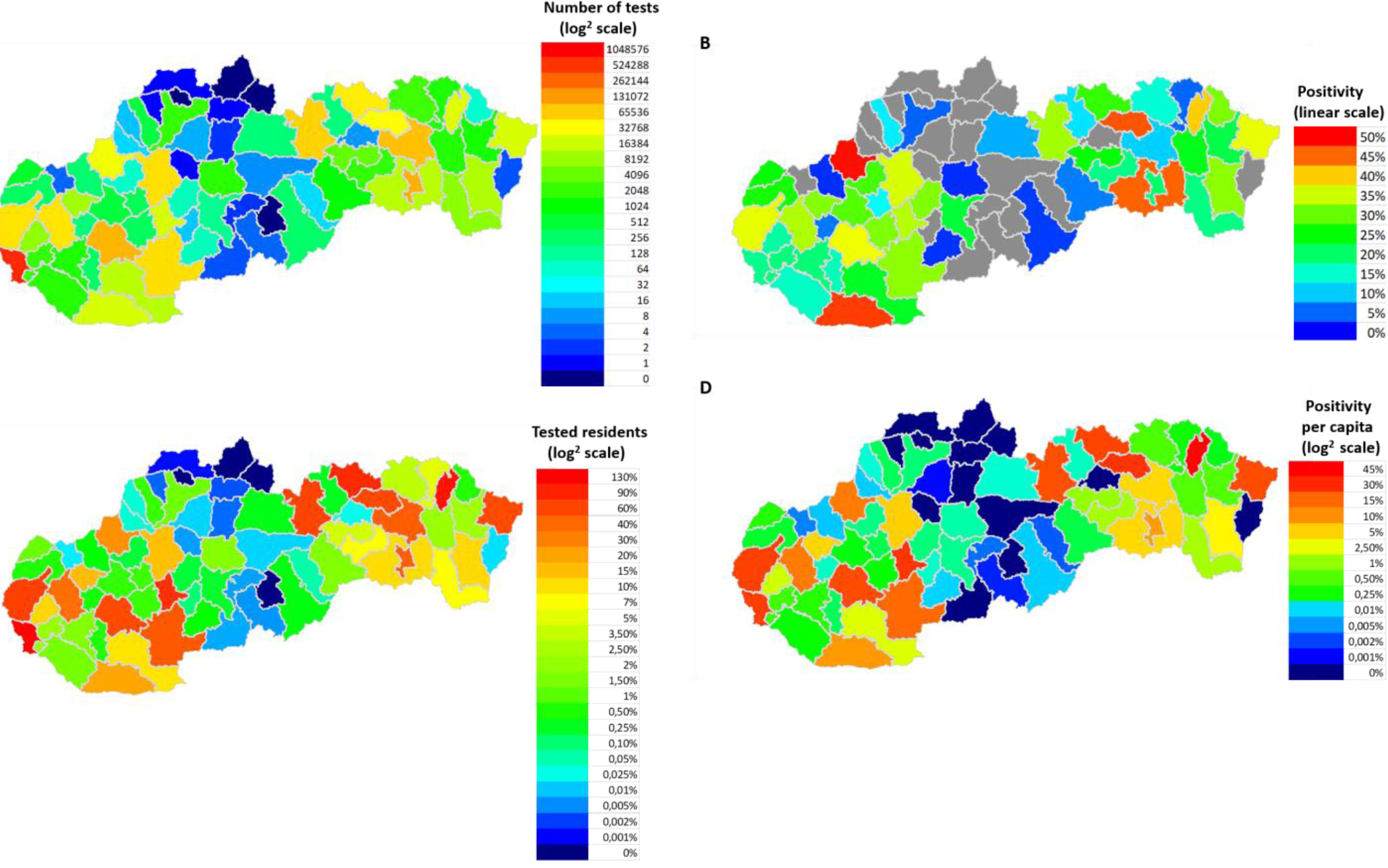
Geographical classification of the examined and positively tested population according to the districts of Slovakia. (A) On the map, each district has a colored representation (log2 scale) corresponding to the number of tests performed. For each color of the scale, the number of tests corresponding to each color is indicated. (B) Each district has a colored representation (linear scale) on the map that corresponds to the total positivity of the district. For each color of the scale, the positivity rate (%) corresponding to the individual colors is given. Non-representative districts with insufficient sample representation (less than 50 samples) to calculate total positivity are shown in gray. (C) Each district has a color representation on the map (log2 scale) that corresponds to the rate of tests performed per district population. For each color of the scale, the rate of frequency (%) corresponding to individual colors is given. (D) Each district has a representation on the map (log2 scale) corresponding to the rate of positive tests for the population of individual districts of Slovakia. For each color of the scale, the positivity rate (%) corresponding to the individual colors is given

**Table 1:** Number of positive/negative tests and total positivity of tests for individual age categories. Age distribution of the tested individuals along with the corresponding positivity rate, calculation of the total positivity of the study tests.

| Age group | Positive | Negative | Total | Positivity (%) |
| --- | --- | --- | --- | --- |
| 0-2 | 1,806 | 9,273 | 11,079 | 16.30% |
| 3-5 | 4,757 | 14,201 | 18,958 | 25.09% |
| 6-8 | 10,689 | 21,112 | 31,801 | 33.61% |
| 9-13 | 26,193 | 42,544 | 68,737 | 38.11% |
| 14-15 | 9,983 | 17,968 | 27,951 | 35.72% |
| 16-19 | 18,839 | 37,543 | 56,382 | 33.41% |
| 20-26 | 28,861 | 90,078 | 118,939 | 24.27% |
| 27-65 | 216,104 | 703,904 | 920,008 | 23.49% |
| >65 | 32,835 | 133,882 | 166,717 | 19.70% |
| **Total** | **350,067** | **1,070,505** | **1,420,572** | **24.64%** |

The most PCR tests were performed in the Bratislava district, their total number was 612,427, which is more than 6 times more compared to the Kosice district (2nd most numerous in tests performed - 100,838 RT-qPCR tests) (Fig. 1A). The rate of the number of tests per population reached 130% in the Bratislava district (Fig. 1C). The highest total positivity (more than 40%) was achieved by the following districts: Sabinov (45.9%), Trencin (49.25%), Komarno (47.01%), Kosice-okolie (45.16%) (Fig. 1B). A more objective view of the rate of positivity in individual districts is represented by the recalculation of the number of positive RT-qPCR tests per district population. In this case, only the district of Stropkov (42.615%) achieved positivity above 40%. (Figure 1D).

The map in Figure 2A visualizes the geographical division of Slovakia into 8 regions - Bratislava, Trnava, Nitra, Trencin, Zilina, Banska Bystrica, Kosice and Presov. The graph of Figure 2B shows the differences in Ct values of positive PCR tests between individual regions. Differences in positivity between all pairs of regions were found to be significant based on the Fisher exact test (p-value < 3.76E-4). A significant difference in Ct values was demonstrated on the basis of the independent samples t-test between all pairs of regions, except for the pairs: BA-TT (p-value 0.99), BA-ZA (p- value 0.14), BA-KE (p-value 0.44), TT-ZA (p-value 0.14), TT-KE (p-value 0.57), NR-ZA (p-value0.7), NR-BB (p-value 0.04), TN-ZA (p-value 0.8), ZA-BB (p-value 0.56), ZA-KE (p-value 0.15), ZA-PO (p-value 0.46), BB-PO (p-value 0.14). The average Ct values of individual regions were equal to values from 26.57 to 27.72.

**Figure 2:**
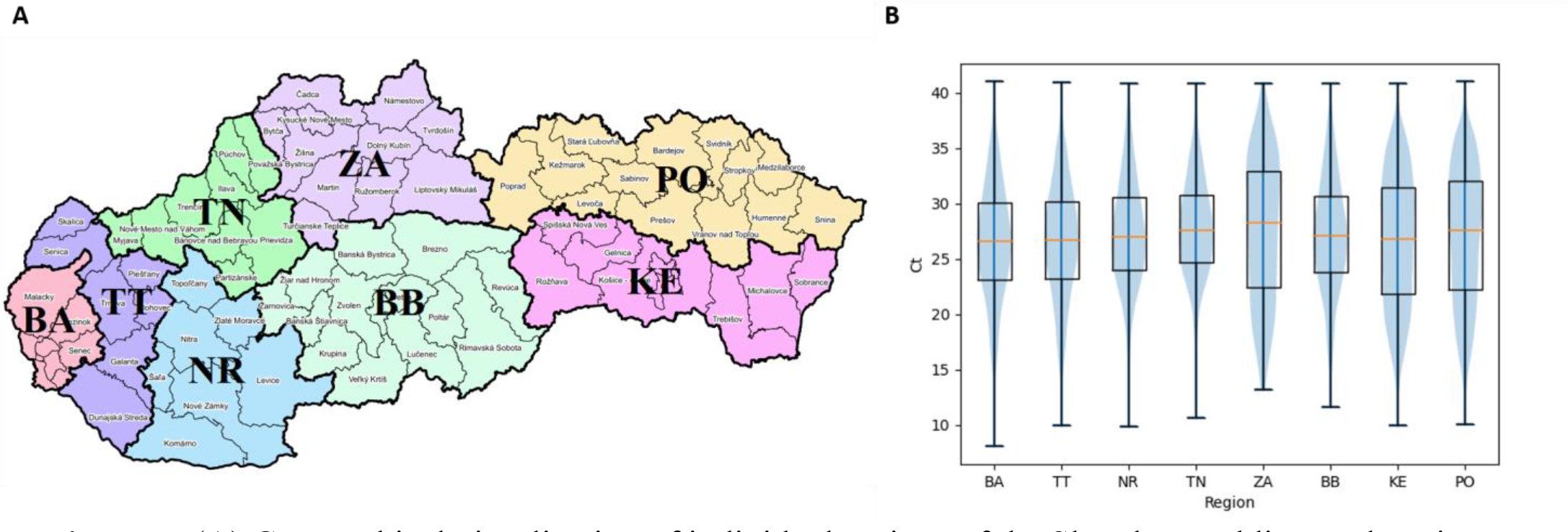
(A) Geographical visualization of individual regions of the Slovak Republic. Explanations: BA - Bratislava region, TT - Trnava region, NR - Nitra region, TN-Trencin region, ZA - Zilina region, BB – Banska Bystrica region, KE - Kosice region, PO - Presov region [17]. (B) Distribution of Ct values of positive diagnostic tests of individual regions of Slovakia shown as boxplots, supplemented by median Ct values (red lines).

The number of tests performed per day with the corresponding number of positive tests is shown in Figure 3. The curve of test frequency and positivity in daily intervals is shown in Figure 3A. For comparison with our data, a similar distribution of frequency and positivity of all performed diagnostic tests within the national testing in Slovakia is shown in Figure 3B. Figure 4 shows the proportion of positivity of tests carried out within the study (4A) and, for comparison, the proportion of positivity of all tests carried out in Slovakia within the framework of national testing (4B). In Figure 4B, the data from March 2020 to September 2022 are plotted with vertical yellow lines. The curves of the graphs show that the positivity rate followed the pandemic waves in Slovakia, which is also represented by the number of tests performed over time (Figure 3, 4).

**Figure 3:**
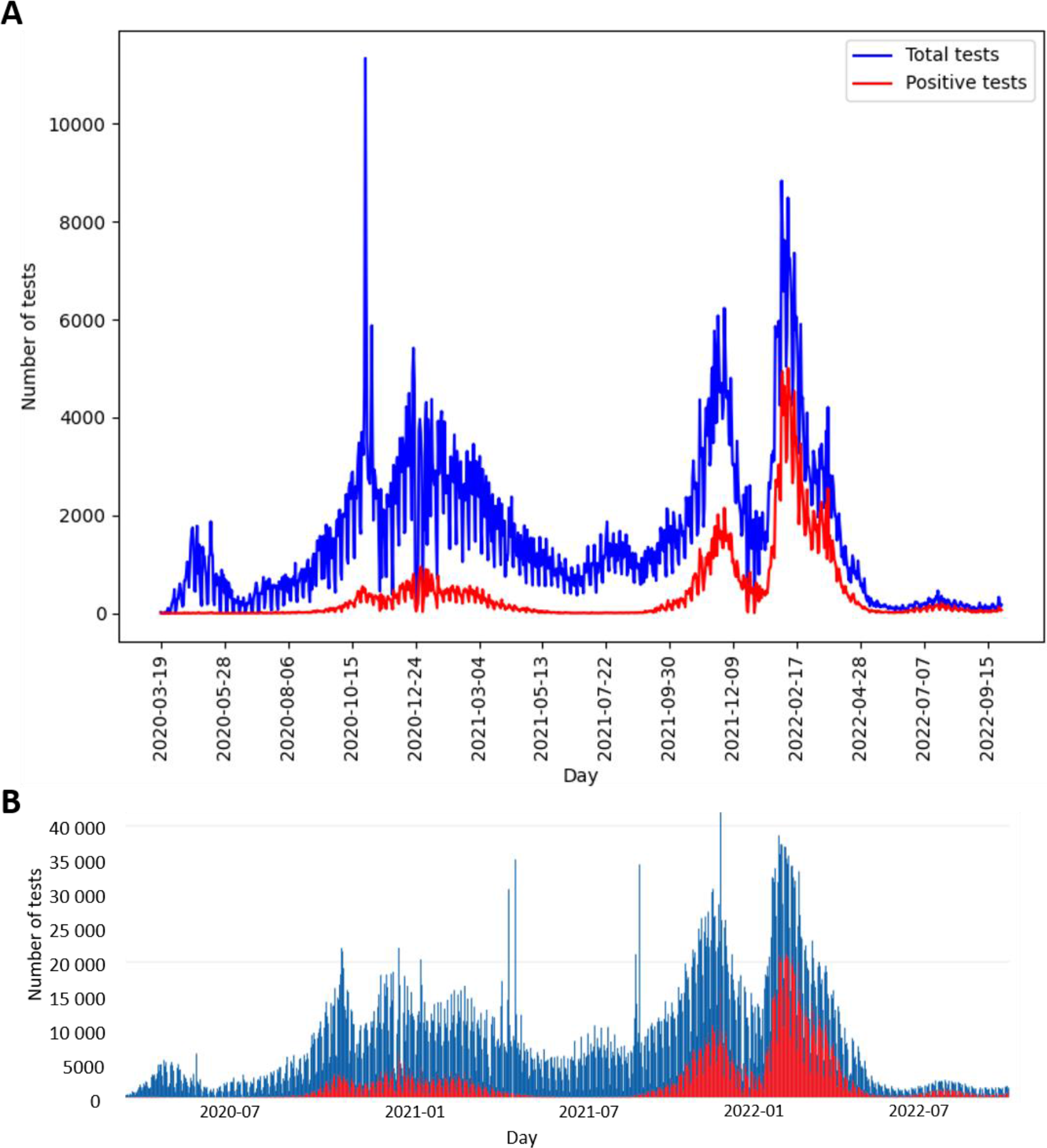
(A) Distribution of study PCR diagnostic tests performed per day (blue curve), corresponding number of positive study tests per day (red curve), compared with (B) distribution of all diagnostic tests performed per day within the Slovakian PCR testing (blue curve), corresponding number of positive tests within the Slovakian PCR testing (red curve) [5]. (A, B) graphs show the date range 2020- 03-19 to 2022-09-29.

**Figure 4:**
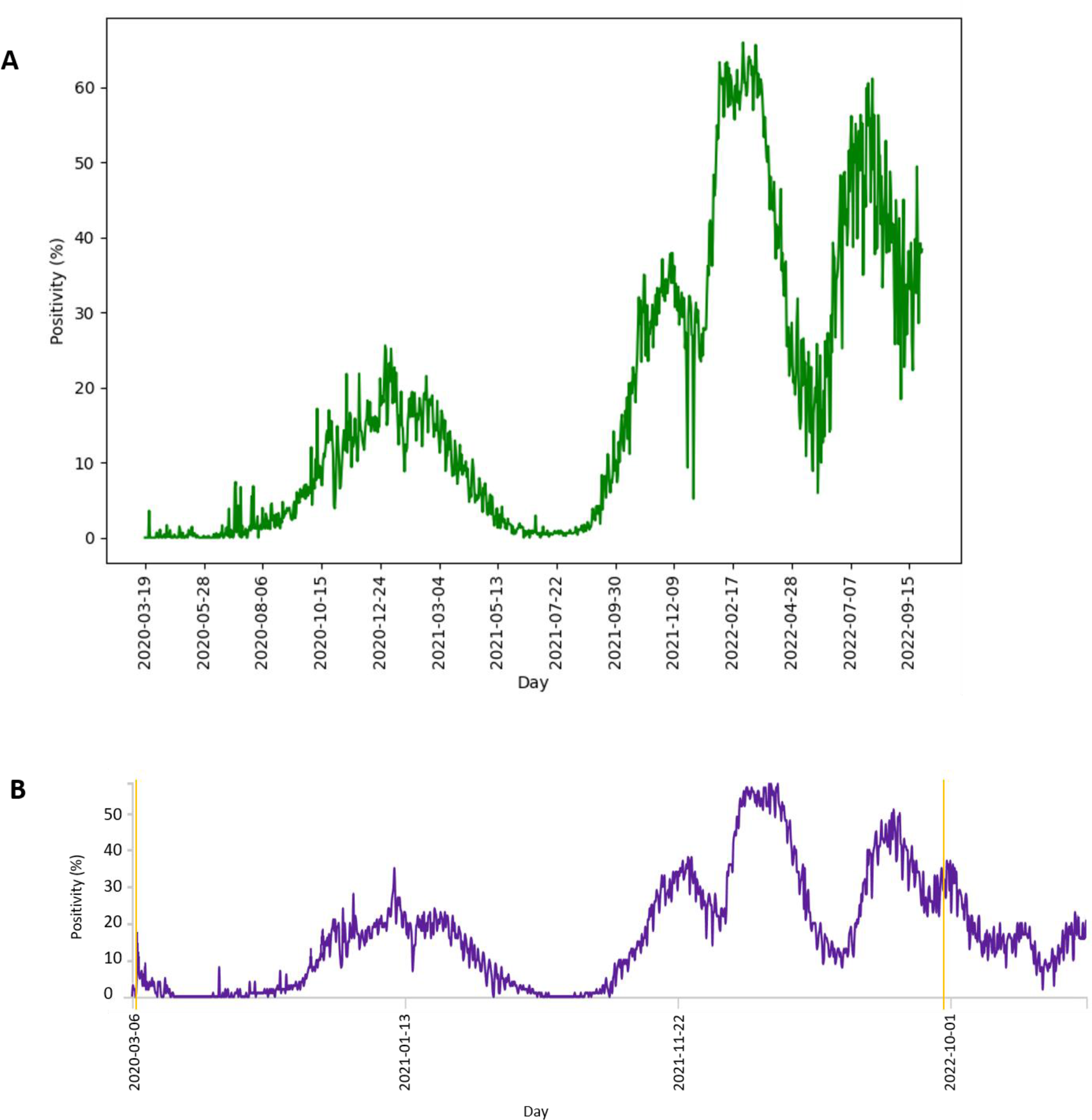
**(A)** Distribution of proportion of positive diagnostic tests of study performed per day, compared with **(B)** distribution of proportion of all positive tests per day within the Slovakian PCR testing [18]. Parallel yellow lines **(B)** define the period from 2020-03-19 to 2022-09-28.

Figure 5 shows the age distribution of tested individuals and positive individuals (5A), respectively the age distribution of tested and positive individuals depending on gender – male/female (5B). Based on the Fisher exact test, the difference between the positivity of men (23.72%) and women (25.54%) was significant (p-value 1.36E-139). The most tested individuals were aged 43 (30,895 individuals). Based on the performed Hartigans’ dip test, the distribution of Ct values of positively tested individuals is probably not unimodal, while the average median of Ct values is equal to 27.9 (5C). The detected Ct values ranged from 9.5 to 41. Based on the independent samples t-test, significant differences in the average Ct values between men and women were demonstrated (p-value 5,88E-27) (5C). The highest average Ct values were achieved in the age group of 3-5 years, equal to the number 30.75, the lowest average Ct values were achieved in the age group > 65 years and represented the number 27 (5D).

**Figure 5:**
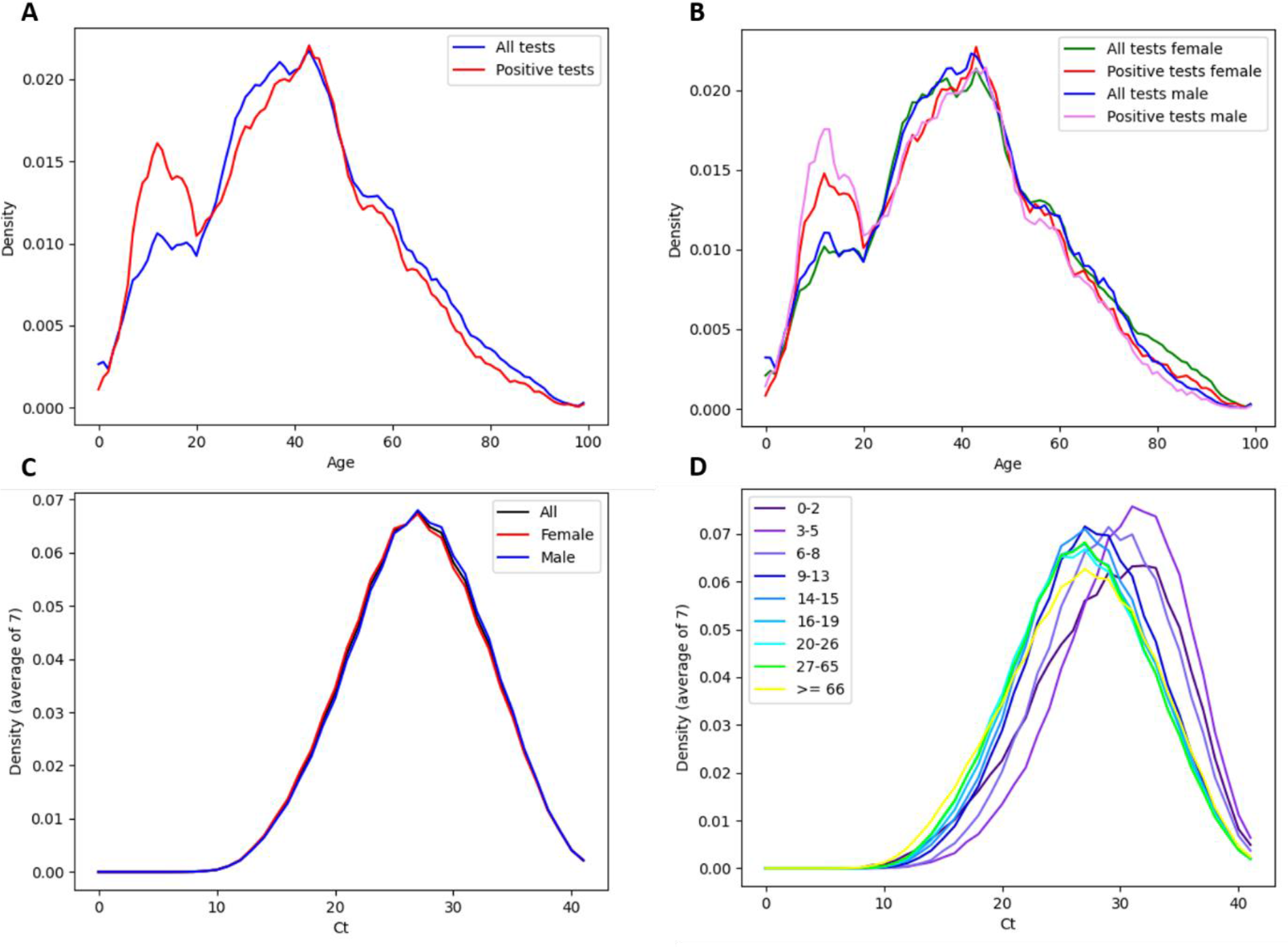
**(A)** Age distribution of all tested individuals (blue curve), positively tested individuals (red curve). (B) Age distribution of all tested women (green curve), positive tested women (red curve), all tested men (blue curve), positive tested men (pink curve). (C) Ct distribution of all diagnostic tests (black curve), tests performed in females (red curve) tests performed in males (blue curve). (D) Ct distribution of all diagnostic tests. The data is broken down by the age of the individuals.

The average weekly Ct of our data reached values from 22.33 (week starting 01/28/2021 - pandemic wave) to 30.12 (week starting 06/30/2022 - pandemic on the wane) (6A). However, the correlation between the change in the number of positives with a one-week delay and the average Ct values based on Spearman’s rank correlation coefficient was not confirmed (p-value 0.52).

Figure 7 shows the distribution of Ct/total positivity values depending on the age of the individuals. The violin chart demonstrates a gradual decrease in the median Ct values along with the increasing age of the categorized study groups. The highest value of the median Ct was found in the age group of 3-5 yrs, equal to the number 30.75. The lowest median Ct value was found in the 20-26 yrs and 27-65 yrs groups, equal to the number 26.7 (7A). Overall positivity above 30% was noted in patients aged 7-19 yrs (7B). A significant difference in positivity between age categories was recorded on the basis of the Fisher exact test between all pairs, with the exception of pairs: 3-5 yrs (positivity 25.09%) and 20-26 yrs (positivity 24.27%) (p-value 0.5); 6-8 yrs (positivity 33.61%) and 16-19 yrs (positivity 33.41%) (p-value 19.88). Significant differences in the average value of Ct within the age categories were demonstrated on the basis of the ndependent samples t-test in almost all pairs with the exception of: 0- 2 yrs and 6-8 yrs (p-value 7.04), 14-15 yrs and 16-19 yrs (p-value 0.2), 20-26 yrs and 27-65 yrs (p-value 33.61), 20-26 y and > 65 yrs (p-value 9.86), 27-65 yrs and > 65 yrs (p-value 6.25).

**Figure 6:**
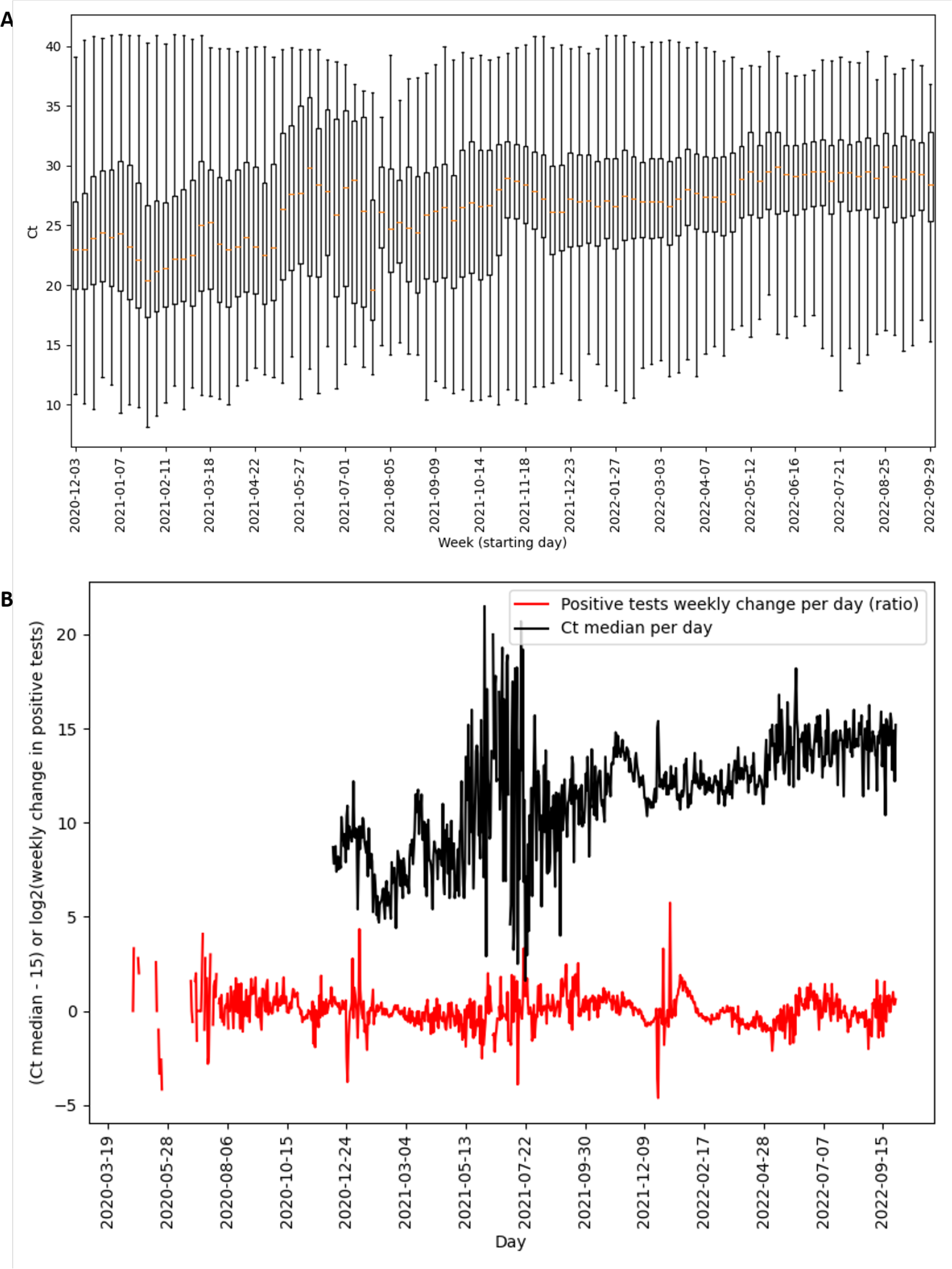
**(A)** Ct distributions of positive diagnostic tests shown as boxplots, supplemented by Ct median values (orange segments). **(B)** Time course of median Ct per day (black curve) and change of ratio posite tests per day with one week delay (red curve). **(A, B)** Gaps in the graphs reconstruct pandemic days/ weeks with an insufficient number of samples (less than 50 performed PCR tests) non- representative results not included in the evaluation.

**Figure 7:**
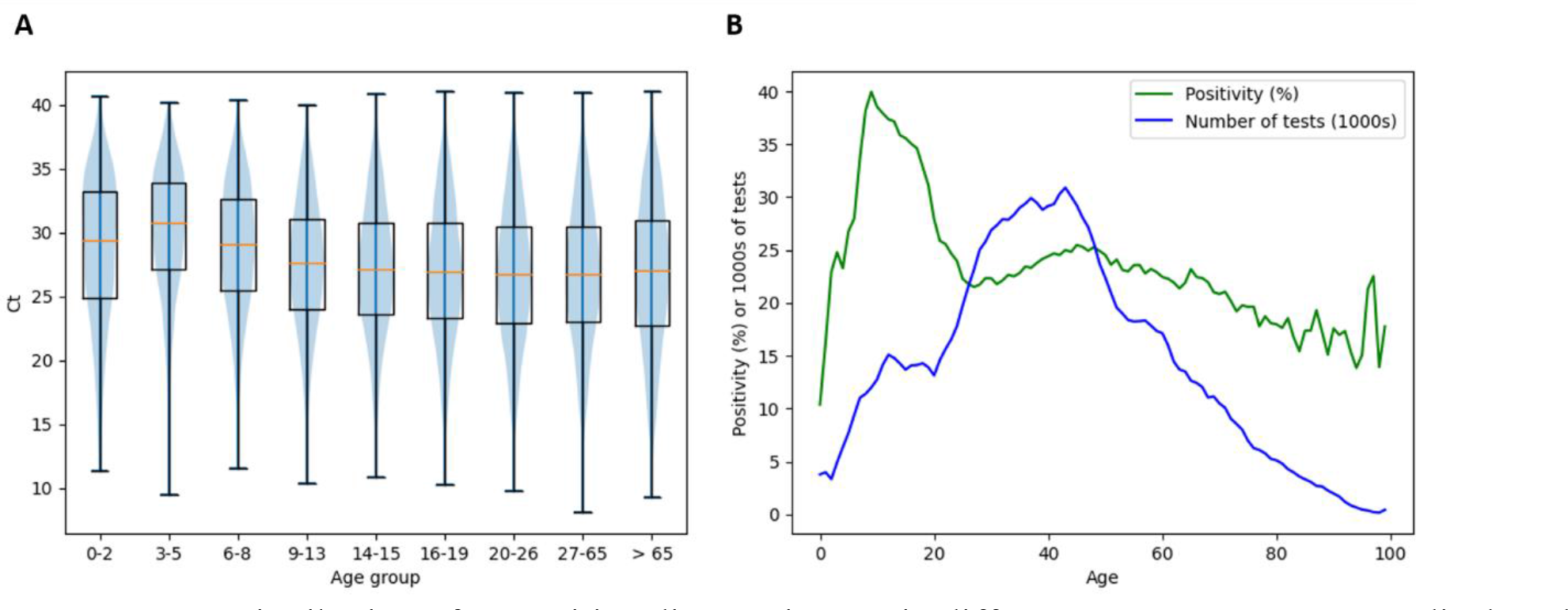
**(A)** Distribution of Ct positive diagnostic tests in different age groups. Data are displayed as violin charts and notched boxplots supplemented with Ct median values (orange segments). **(B)** Distribution of positivity rate by age of individuals (green curve) and distribution of number of tests by age of individuals (blue curve).

### 3.3 Discussion

The COVID-19 pandemic has affected many areas of medicine and healthcare around the world. Its further development is very difficult to predict. The situation is complicated by the fact that the virus is able to constantly evolve and generate new mutations, which in a specific constellation can lead to the emergence of new variants that can have different manifestations (infectivity, mortality). The inability to predict increases or decreases in cases of SARS-CoV-2 infection has significantly affected the ability of public health officials to respond to this crisis and take the necessary measures. Early detection of the SARS-CoV-2 virus using diagnostic methods is therefore still crucial. The optimal method continues to be the RT-qPCR method, which provides a high diagnostic potential thanks to the currently well-developed laboratory infrastructure.

In our study, we analyzed whether and how average weekly Ct values from RT-qPCR tests correlate with the occurrence of SARS-CoV-2 infection in a subpopulation of Slovak residents tested in one accredited laboratory, the tested individuals were classified into age groups. The actual local prevalence and the number of tests performed affect the value of the positivity rate. In general, as more tests are deployed, the positivity rate will approach the true prevalence [19]. The analyzed tests of this study represent > 19% of all diagnostic PCR tests in Slovakia (for the given date interval) from > 93% of the districts of the Slovak Republic, so they should be representative of the population, which is also shown by comparing the frequency and positivity curves of the tests in our study with the curves of the data of the national Slovakia’s Covid-19 PCR testing. Although the data showed that children in primary and secondary schools (6-19 years old) had the highest positivity rate (Table 1, Figure 7B), this phenomenon is probably related to the lower frequency of tests in the given age subgroup. In addition, children had schools closed during much of the pandemic [20] and were not required to undergo regular PCR testing (e.g. mandatory for some groups of employees in the workplace). Therefore, children with symptoms of the disease apparently requested PCR diagnosis of SARS-CoV- 2 as a priority, which could also have contributed to a higher rate of overall positivity in their age subgroup.

The number of performed RT-qPCR tests in Slovakia was largely influenced by changes in the availability of a free RT-qPCR test. From May 2022, a referral from a doctor (general practitioner or specialist) is required to perform a free RT-qPCR test. The exception is patients over 60 years of age, who can continue to request a free RT-qPCR examination through the state portal [21] in case of symptoms of the disease. In the aforementioned study, a lower frequency of performed tests is detected in the time interval April/May 2022. From mid-May 2022 until the end date of the analyzed data of the study, which is September 28, 2022, a significantly reduced frequency of tests was subsequently detected (Figure 3A). A possible limitation in the geographical (districts of Slovakia) classification of performed and positively evaluated PCR tests could be the migration of residents (e.g. due to employment) within districts without updating the permanent residence of citizens. In the result of our study, this was confirmed by recalculating the number of tests performed in the Bratislava district to the official number of inhabitants of this district - probably for this reason, the ratio of the number of tests to the number of inhabitants of the Bratislava district reached up to 130%.

The results of several studies show that the distributions of Ct values in SARS-CoV-2 screening cases are typically bimodal [22, 23]. There is an assumption that a lower maximum Ct value corresponds to patients with high infectivity and a higher maximum Ct value corresponds to patients with low infectivity [23]. A bimodal distribution is also typical for other viral infections, with the lowest Ct values indicating the proportion of individuals with a high viral load in the acute phase of infection, and higher Ct values being typical for the early phase of infection or convalescence [11, 24, 25]. This assumption could explain why the distribution of average Ct values of our data is not unimodal.

There are also currently several studies determining the ability of Ct values to predict future cases of COVID-19 in terms of outcomes [26, 27, 28]. Although in several published studies, summary analyzes of the distribution of Ct values revealed that the average weekly Ct values fluctuated during the pandemic and that a gradual decrease in the average weekly Ct values indicated a future increase in the occurrence of positive samples [29], this trend was not confirmed by the results of the analysis on our data. Most relevant potential reasons for this difference may be the chaotic organization of the screening and diagnostics in Slovakia performed by the laboratories and relatively unspecific and broadly described criteria regarding the public testing. As in our dataset no specific selection of samples was done, e.g. positive for specific symptoms or based on origin related to specific organizations indicating the test (hospitals, ambulances, selected collection points) the data could be not able to be used for such evaluation and potential prognostic use.

Next, we focused on the distribution of Ct values in different age groups to determine which groups may contribute the most to the spread of infection. Biomedical research today relies on p-values as a deterministic measure for data-driven decision making, where a single p-value is calculated from samples of data to determine statistically significant differences between groups of observations. Because the estimated p-value tends to decrease with increasing sample size, applying this methodology to data sets with large sample sizes leads to a higher probability of rejecting the null hypothesis [30], which was also reflected in our data (more than 1.4 million RT-qPCR tests). This fact probably contributed to the demonstration of significant differences in the average Ct values between almost all categorized age groups and also between the regions of Slovakia. This assumption is also confirmed by the fact that the differences between the average Ct values of the Zilina region, which has the smallest number of samples (95 RT-qPCR tests), and the remaining regions were not significant in any case. However, this does not mean that the observed significance is not real. On the contrary, low p-values are present with large sample sizes because the robustness of data means that even a small difference is more likely to reflect a real underlying difference between two datasets. We observe many significant differences between various groups of patients and it can be explained by high complexity of the pandemic. Although there were general trends followed by most patient groups, too many variables participated in the outcome. Examples include infection outbreaks being present in different locations at different times or high variability of measures for suppressing the pandemic.

However, the graph in Figure 7A demonstrates that the decrease in mean Ct values was consistent with increasing age of individuals in age-related subgroups. This observation is consistent with the statement that children with SARS-CoV-2 infection are often asymptomatic or mildly symptomatic and are rarely the index case in household transmission chains [31]. There are several factors that can potentially affect the detected Ct values, including the type of sample (e.g. nasopharyngeal, anterior nasal mucosa, saliva, sputum) or the efficiency of sampling, (non-intensive sampling can lead to inaccuracies of the results) [32]. The detection of lower Ct values in children may be related to the quality of sampling, as it is necessary to introduce the sampling rod into the nasopharynx [33]. Saliva sampling was put into practice precisely because of the complications of sampling from the nasopharynx in certain groups of individuals - significant pain during nasopharyngeal swabs was described by 58% of young people compared to none during saliva sampling, while 90% of children prefer saliva sampling [34]. The results of the articles demonstrate that detection from saliva samples performs as well or better than nasopharyngeal swabs in hospital, emergency care and mass screening settings for SARS-CoV-2 testing by RT-qPCR [35–38]. However, the average Ct value in saliva can be higher compared to the average Ct value from nasopharyngeal swabs [39]. Therefore, a combined nasal and throat swab is usually recommended as a more suitable sample for RT-qPCR diagnosis [32]. As we wanted to include in this study all samples from March 2020 - September 2022 provided by the Medirex laboratory with a positive/negative result detected, the data includes both nasopharyngeal swabs and saliva samples. However, the results representing the detection of SARS-CoV-2 from saliva samples represent less than 5% of all samples and therefore we do not assume that this fact could affect the overall informative value of the average Ct values for the data of this study.

### 3.4 Conclusions

The age of patients was the most relevant individual parameter when it comes to differences in the viral load as clinically relevant and patient cohort characteristics. The lower age was associated with higher Ct values, and so with lower viral load, and decreased with the age increase. This finding could be used in the public services or lockdown planning during pandemic, e.g. to let the schools open could be less risky when it comes to disease spread in comparison to other public services.

However, it is important to noted that the decision to introduce, adapt or cancel public health and social measures should be based primarily on a situational assessment of the intensity of transmission, the vaccination rate of the population and the ability of the health system to respond to a pandemic.

## Data Availability

All data produced in the present study are available upon reasonable request to the authors

## 5 Ethics statement

The study was conducted in accordance with the Declaration of Helsinki, and approved by the Ethical committee of Bratislava Self-Governing District under the identifier 03228/2021/HF from January 12, 2021. All patients have filled the questionnaires with relevant information regarding their health status in relation to COVID-19 and signed informed consent.

## 7 Author Contributions

Conceptualization, GM and VR; methodology, GM, AG and NJ; software, AG; validation, GM and NJ; formal analysis, AG; investigation, DK, GB and RS; resources, EM and ET; data curation, EM and ET; writing—original draft preparation, NJ; writing—review and editing, GM; visualization, NJ, GM and AG; supervision, GM; project administration, GM; funding acquisition, GM and VR. All authors have read and agreed to the published version of the manuscript. All authors contributed to the article and approved the submitted version.

## Acknowledgments

The authors thank the laboratory Medirex, a.s. for providing data for the study.

## 10 Supplementary Material

The following supporting information - table with all samples and laboratory information included in the analysis can be downloaded at:

## 11 Data Availability Statement

NA.

